# Convergent connectivity of sleep disorders and its neurotransmitter and cell enrichment correlates

**DOI:** 10.64898/2026.09.09.26362596

**Authors:** Mortaza Afshani, Amin Saberi, Gerion M. Reimann, Congying Chu, David Elmenhorst, Leon Lotter, Sofie L. Valk, Juergen Dukart, Sarah Genon, Veronika I. Müller, Simon B. Eickhoff, Masoud Tahmasian

## Abstract

Sleep disorders are highly prevalent and impose a substantial public health burden, yet the neurobiological mechanisms underlying shared and disorder-specific abnormalities remain incompletely understood. We conducted a pre-registered transdiagnostic study to identify shared and distinct normative macroscale connectivity patterns associated with reported brain abnormalities across insomnia disorder, obstructive sleep apnea, narcolepsy, and rapid eye movement sleep behavior disorder, and to characterize their microscale neurotransmitter and cellular correlates. We obtained peak coordinates from 80 experiments reporting meta-analytic brain abnormalities across these four sleep disorders. Convergent connectivity mapping was then applied to these coordinates using dense connectivity matrices derived from 100 unrelated participants from the Human Connectome Project, with connectivity patterns evaluated relative to a null distribution generated from randomly selected foci. We subsequently examined the spatial correspondence of the identified connectivity maps with 19 neurotransmitter systems using curated positron emission tomography-derived density maps and assessed cellular enrichment across 24 cell-type systems using regional gene-expression profiles from the Allen Human Brain Atlas. Statistical significance was assessed using spin-based permutation testing and false discovery rate correction (pFDR < 0.05). Shared convergent connectivity patterns were identified within the default mode network (zmean = 1.103, pspin,FDR = 0.019), as well as the dorsal attention (zmean = −0.674, pspin,FDR = 0.014) and frontoparietal (zmean = −0.239, pspin,FDR = 0.019) resting-state networks. Disorder-specific analyses revealed broadly overlapping connectivity patterns, with some disorder-related variability. Both transdiagnostic and disorder-specific connectivity patterns showed selective spatial correspondence with multiple neurotransmitter systems, including serotonin, dopamine, norepinephrine, and glutamate, and were enriched for excitatory, inhibitory, and non-neuronal cell types (pFDR < 0.05). These findings indicate that sleep disorders share convergent macroscale connectivity architecture while also exhibiting disorder-specific variations, with the identified network patterns linked to distinct neurotransmitter and cellular profiles. Together, the results provide a normative systems-level framework for understanding shared and distinct neurobiological mechanisms across sleep disorders.

## Introduction

Chronic sleep disorders, including insomnia disorder (ID), obstructive sleep apnea (OSA), narcolepsy, and rapid eye movement sleep behavior disorder (RBD), are prevalent worldwide^1–4^ and impose a substantial burden on global public health.^5,6^ They are risk factors for, or comorbidities of, neuropsychiatric conditions and increase the risk of mortality.^7–12^ Understanding their neurobiology is therefore essential for improving diagnosis and treatment. Neuroimaging studies have identified structural and functional brain alterations and provided insights into their cellular and molecular substrates^13–17^, but their multiscale neurobiological mechanisms remain poorly understood.

While sleep disorders have distinct etiologies and clinical characteristics, they share substantial symptom and biological overlap. Common features include poor sleep satisfaction and efficiency, sleep fragmentation, daytime sleepiness, cognitive impairment, emotion dysregulation, and mood disturbances.^18–20^ In addition, large-scale genome-wide association studies identified shared polymorphisms, overlapping genomic loci, and biological pathways across multiple sleep-related symptoms.^21,22^ The presence of overlapping symptom profiles, homologous comorbidities (e.g., depression), and shared genetic architectures call for a transdiagnostic approach to identify their shared abnormalities.

Our recent coordinate-based meta-analysis (CBMA) across several sleep disorders identified convergent regional abnormalities in the bilateral subgenual anterior cingulate cortex (sgACC), the right amygdala, and hippocampus.^15^ CBMA enables quantitative synthesis of neuroimaging findings by identifying brain regions consistently associated with specific cognitive functions or disorders.^23,24^ However, it is designed to identify regional convergence of effects and therefore does not capture connectivity between affected brain regions or the broader networks they comprise. To address this limitation, *convergent connectivity mapping (CCM)*^25,26^ has been proposed to assess network-level convergence of effects by comparing the normative connectivity profiles of reported coordinates in prior studies with those of randomly sampled coordinates. Applying this approach to depression has demonstrated that, even in the absence of regional convergence^27^, aberrant brain regions converge at the network-level^25^, aligning with clinically meaningful circuit-level models. Sleep disorders are similarly recognized as disturbances of interacting large-scale brain networks, with abnormalities distributed across the whole brain.^16,28,29^ Yet, while our CBMA^15^ revealed consistent regional abnormalities across several brain areas, it remains unclear whether these findings converge at the network level within specific circuits. Applying CCM to decades of neuroimaging research can therefore provide a systems-level characterization of distributed abnormalities, moving beyond regional localization toward an integrated understanding of brain dysfunction.

Macroscale brain abnormalities in sleep disorders are shaped by underlying molecular and cellular mechanisms, highlighting the need to examine macro–microscale interactions. Molecular imaging suggests that imbalances in neurotransmitter systems, including dopamine, norepinephrine, serotonin, GABA, and glutamate, may disrupt sleep–wake regulation and increase susceptibility to sleep disorder^30–32^. Animal models further suggest that sleep loss is associated with altered expression of genes involved in synaptic signaling, as well as region-specific transcriptional reprogramming in non-neuronal cell types, which can lead to sleep dysregulation and circadian dysrhythmia.^33–35^ Nevertheless, how these molecular and cellular alterations relate to macroscale brain abnormalities across chronic sleep disorders remains unclear.

In this pre-registered (https://osf.io/afvp8) study, we aimed to address these questions: *i) Do brain regions with aberrant structure or function across sleep disorders exhibit shared and/or disorder-specific macroscale convergent connectivity patterns at the network level? ii) Are there transdiagnostic and disorder-specific microscale neurotransmitter and transcriptomic correlates of the identified convergent connectivity maps?* Thus, we employed sleep disorder-related peak coordinates from our prior meta-analysis^15^ and calculated transdiagnostic and disorder-specific CCMs. We then performed spatial correlations between these CCMs, 19 neurotransmitter systems, and 24 cell-type systems using transcriptomic data.

## Methods

### Sleep disorders meta-analytical data

We obtained the peak coordinates of brain alterations associated with different sleep disorders from our prior CBMA^15^. We included data from 80 experiments (i.e., individual group-level contrasts comparing a specific functional or structural neuroimaging marker between patients and control groups), comprising 1,146 stereotactic coordinates of significant clusters and 1,937 participants. We selected only disorders with more than 10 reported experiments, resulting in the inclusion of four sleep disorders: ID, OSA, narcolepsy, and RBD.

### Resting-state fMRI preprocessing and dense functional connectivity computation

Resting-state fMRI data were obtained from 100 unrelated Human Connectome Project participants (mean ± SD age: 29.11 ± 3.68; 54% female). Each participant completed four scanning sessions (14:33 min/session; TR = 0.72 s; acquisition details elsewhere^36^). For each session, minimally processed MNI-space data were preprocessed using nilearn, including detrending, temporal band-pass filtering (0.01–0.08 Hz), spatial smoothing (6 mm FWHM Gaussian kernel), voxel-wise z-scoring, and gray matter masking (>10% probability threshold; 211,590 total voxels). CCM analyses were conducted at the parcel level, excluding non-gray matter voxels. We concatenated preprocessed data across the four sessions per subject. For each participant, Pearson correlation coefficients were computed between BOLD time series for all gray matter voxel pairs, yielding individual dense resting-state functional connectivity (RSFC) matrices. Coefficients were Fisher Z-transformed and capped between -4 and 4 to limit extreme values. To enable efficient large-scale computation, calculations were GPU-parallelized using CuPy. Finally, subject-level dense RSFC matrices were averaged across all 100 participants to generate a group-averaged matrix for subsequent CCM analyses^25,26^. See Supplement for details.

### Convergent connectivity mapping analysis

Using healthy individual connectivity data, we performed CCM to identify convergent connectivity patterns across sleep disorder experiments compared with randomly distributed foci. Peak coordinates were mapped to the nearest gray matter voxel (excluding foci >4 mm away) to extract RSFC profiles from the group-averaged dense matrix. Profiles were weighted by sample size and averaged across foci within experiments to yield an observed RSFC map, which was parcellated using the Schaefer 400 cortical and Tian subcortical (scale S2) atlases. This true map was compared against a null distribution created from 1,000 random-foci permutations using identical two-level averaging and parcellation. Parcel-wise one-tailed p-values reflected the proportion of null iterations equaling or exceeding empirical values. See Supplement for details. We conducted CCM analyses across experiment subsets, including transdiagnostic, disorder-specific, and modality-specific analyses. To assess whether CCM effects were preferentially anchored within the seven canonical resting-state networks^37^ beyond intrinsic spatial smoothness, we performed spatial spin permutation testing by rotating observed maps 1,000 times across the cortical surface, preserving spatial autocorrelation and hemispheric topology. See **Figure 1** and Supplement for details.

**Figure 1.**
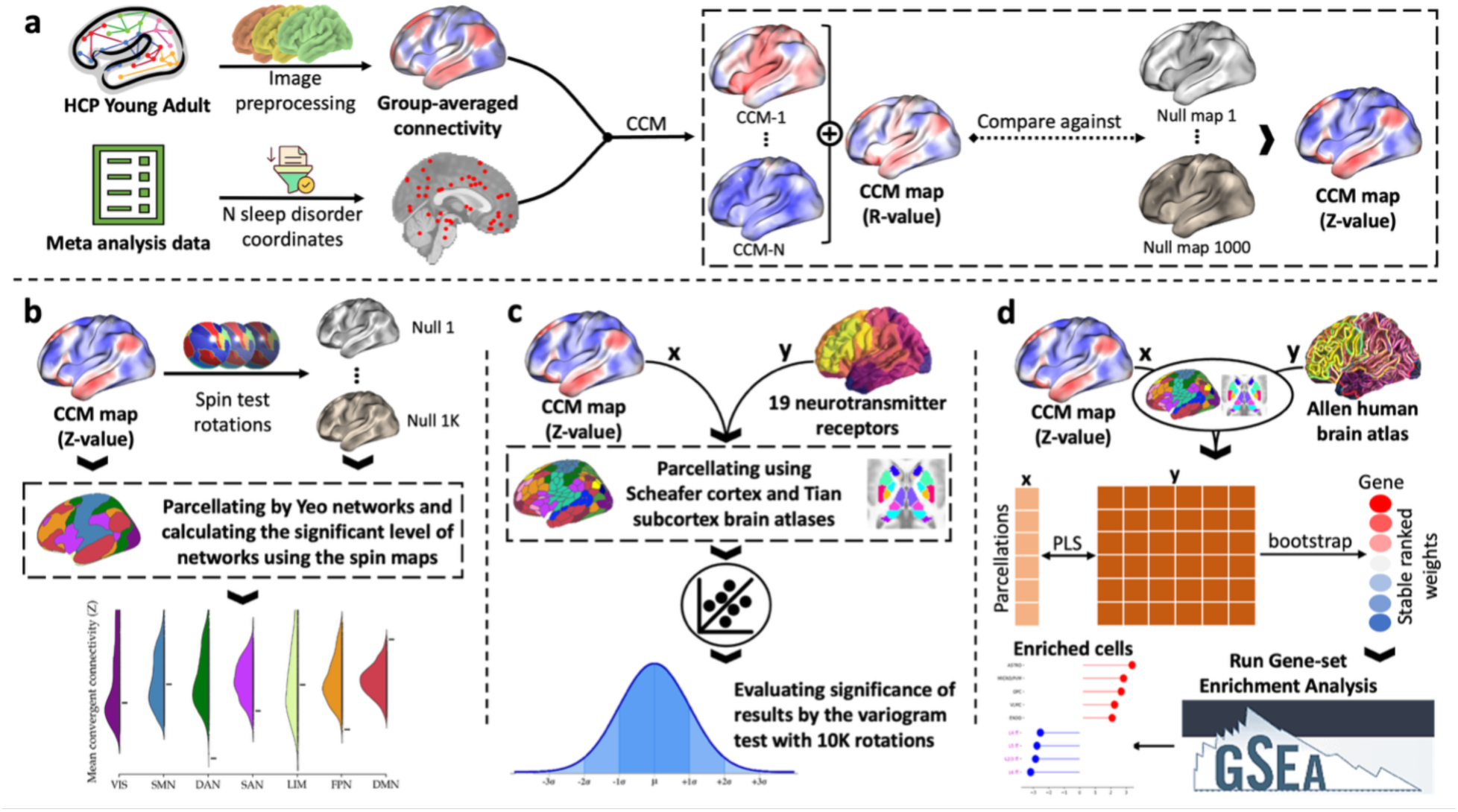
Convergent connectivity mapping and multiscale contextualization of sleep disorder–related brain alterations. **(a)** Peak coordinates from a prior meta-analysis of insomnia disorder, obstructive sleep apnea, narcolepsy, and REM sleep behavior disorder were used to perform convergent connectivity mapping (CCM). A group-averaged dense resting-state functional connectivity matrix derived from the Human Connectome Project was used to estimate the normative connectivity profile of each focus. Connectivity profiles were averaged within and across experiments (sample-size weighted) and compared with randomly sampled foci to generate CCM z-maps. Transdiagnostic and disorder-specific analyses were conducted separately. **(b)** Network-level enrichment of CCM effects was evaluated within the canonical resting-state networks defined by Yeo using spatial spin permutations. **(c)** Neurotransmitter contextualization was performed by correlating CCM maps with PET-derived neurotransmitter receptor and transporter maps parcellated using the Schaefer Atlas and Melbourne Subcortical Atlas. Statistical significance was assessed using spatial autocorrelation–preserving permutations with false discovery rate correction. **(d)** Transcriptomic associations were examined using gene-expression data from the Allen Human Brain Atlas. Partial least squares regression identified gene sets associated with CCM patterns, and Gene Set Enrichment Analysis was used to identify enriched cellular systems, with multiple-comparison correction applied.

### Neurosynth and connectivity colocalization

To evaluate cognitive processes that colocalize with our identified CCM maps, we obtained probabilistic meta-analytic maps for 123 cognitive terms from Neurosynth. These maps were parcellated using the Schaefer 400 and Tian S2 atlases. We quantified spatial correspondence between parcellated CCM maps and meta-analytic cognitive terms using Spearman’s rank correlations across all parcels, and visualized top-ranking terms via word clouds. See Supplement for details.

### Neurotransmitter and connectivity colocalization

To assess the association between the identified connectivity networks (CCM values of transdiagnostic and disorder-specific maps) and neurotransmitter systems, we performed a neurotransmitter contextualization analysis. Here, we correlated Positron Emission Tomography (PET) maps for 11 neurotransmitter receptor/transporter families (serotonin, nicotinic, cannabinoid, dopamine, GABA, histamine, muscarinic, opioid, norepinephrine, glutamate, cholinergic)^38^ with the CCM values derived from the transdiagnostic and individual sleep disorder analyses. The maps were parcellated using the Schaefer-400^39^ and Melbourne S2^40^ atlases and then normalized by Z-scoring. When multiple neurotransmitter maps existed, a sample-size-weighted average was used. To account for spatial autocorrelation, we assessed the statistical significance of correlations against a null distribution generated via variogram-based permutations of the neurotransmitter maps. All p-values were corrected for multiple comparisons using false discovery rate (FDR) adjustment (p_variogram, FDR_ < 0.05). See Supplement for details.

### Cellular gene set enrichment analysis

To assess the link between CCM values and transcriptomic profiles, gene-expression data from six post-mortem brains in the Allen Human Brain Atlas were processed using the abagen toolbox ^41^. Tissue samples were mapped to a combined atlas of 432 regions (400 Schaefer cortical and 32 Melbourne subcortical parcels). Samples were assigned within 2 mm, mirrored bilaterally, and missing data were interpolated using distance-weighted averages. Values were normalized within each donor across genes and samples. After gene stability analysis, we averaged samples across donors into expression matrices for 12,506 genes. We performed Partial Least Squares (PLS) regression to relate gene expression patterns to CCM values. Gene stability for the first PLS component was assessed via 10,000 bootstrap iterations. Stable gene sets were analyzed using Gene Set Enrichment Analysis (GSEA) to identify enriched cellular systems in a single-cell RNA sequencing dataset^42^, with results adjusted for multiple comparisons using FDR correction (p_FDR_ < 0.05). See Supplement for details.

## Results

### Transdiagnostic (shared) convergent connectivity maps

We calculated z-scores of CCM maps, indicating how strongly brain areas are functionally connected (based on a normative connectome) to the reported foci of sleep abnormalities, compared with null connectivity maps generated from random foci. The transdiagnostic CCM map, compared to the null maps, revealed relatively stronger connectivity in cortical areas such as posterior cingulate cortex and ACC, frontal medial cortex, precuneus, and parahippocampal areas, as well as in subcortical regions, including the nucleus accumbens, amygdala, and hippocampus, whereas other brain regions such as supramarginal gyrus exhibited lower connectivity to the foci (**Figure 2A**). We then assessed the co-localization of the transdiagnostic CCM map with the canonical resting-state networks^37^, by comparing the average CCM values within each network against a null distribution generated using spin permutation. The CCM values were significantly lower than expected by chance in the dorsal attention (z_mean =_ -0.674, p_spin, FDR =_ 0.014) and frontoparietal networks (z_mean =_ -0.239, p_spin, FDR =_ 0.019) and were significantly higher in the default mode network (DMN) (z_mean =_ 1.103, p_spin, FDR =_ 0.019) network (**Figure 2B**). This indicates that the reported foci of abnormalities in sleep disorders tend to be connected to, or located within, the DMN, while showing the opposite tendency with respect to the dorsal attention and frontoparietal networks.

**Figure 2.**
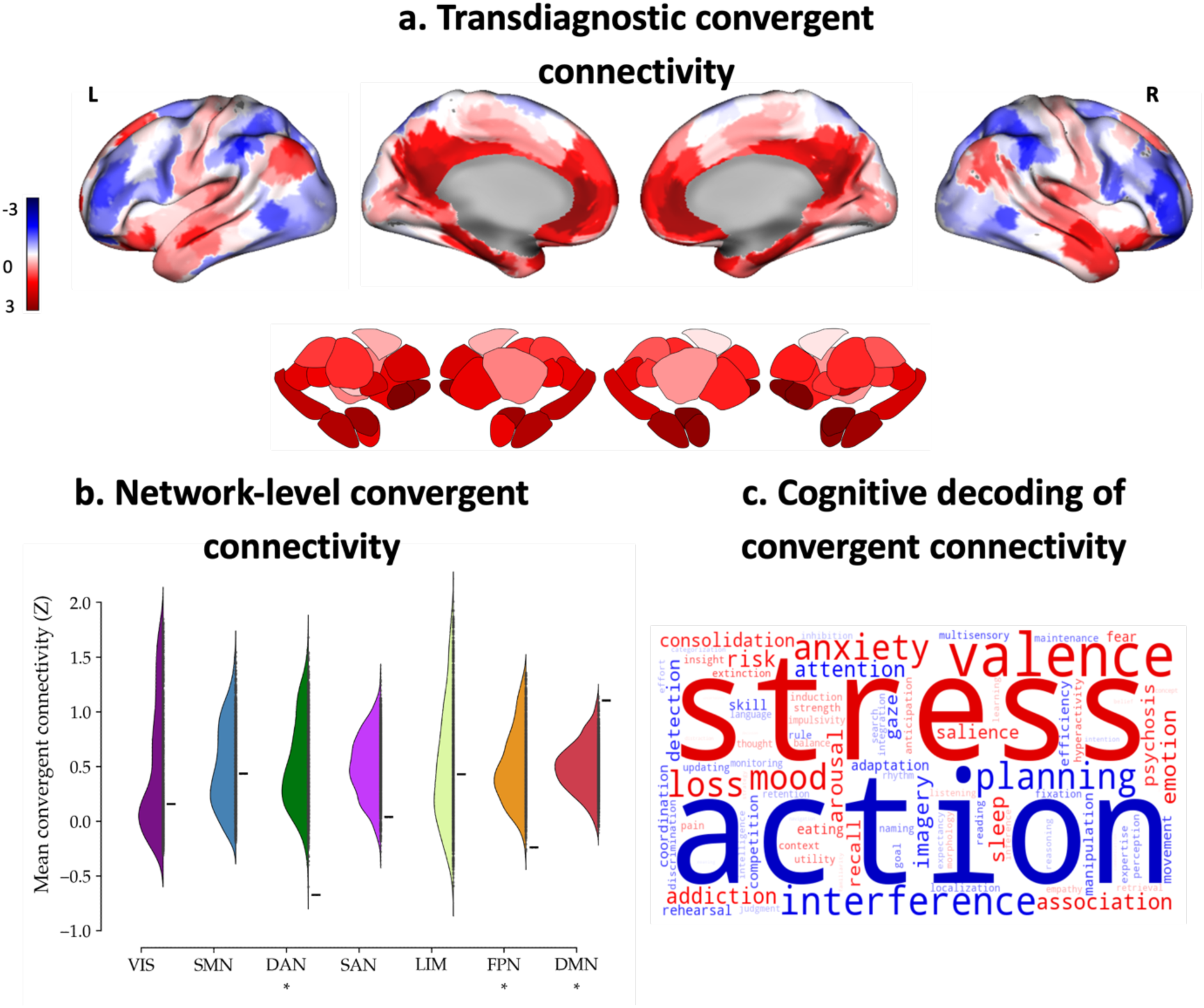
Transdiagnostic convergent connectivity mapping across sleep disorders. **a)** Showing elevated connectivity between the coordinates and subcortical regions (caudate, putamen, amygdala), alongside associated functional circuits, while most remaining brain regions exhibited lower connectivity to the foci. **(b)** Testing the significance of altered mean spatial association with canonical resting-state networks against a null mean derived from random foci revealed that CCM values within dorsal attention and frontoparietal networks were significantly lower, while CCM values for the default mode network were significantly higher. **c)** Cognitive term decoding via spatial correlation with meta-analytic functional maps from the Neurosynth database illustrates the top behavioral and cognitive terms associated with the shared topography, where font size reflects relative association strength. Asterisks denote P_spin, FDR_ < 0.05. *VIS visual network, SMN somatomotor network, DAN dorsal attention network, SAN salience network, LIM limbic network, FPN frontoparietal network, DMN default mode network*.

### Disorder-specific convergent connectivity maps

After characterizing the shared convergent connectivity of sleep disorder abnormalities, disease-specific CCM analyses were performed. We applied the CCM method separately to 26 ID experiments (284 coordinates; 794 participants), 25 OSA experiments (485 coordinates; 583 participants), 16 RBD experiments (199 coordinates; 325 participants), and 13 narcolepsy experiments (178 coordinates; 235 participants). Across disorders, disease-specific reported foci of brain abnormalities showed distinct normative connectivity profiles within functional networks. Coordinates associated with ID showed stronger connectivity with the Planum Polare, Cingulate Gyrus (anterior division), and Supplementary Motor Cortex, alongside weaker connectivity with the Angular Gyrus, Frontal Pole, and Middle Frontal Gyrus. Coordinates for OSA demonstrated stronger connectivity with the Precuneus, Middle Temporal Gyrus (anterior division), and Cingulate Gyrus (posterior division), but weaker connectivity with the Supramarginal Gyrus (anterior division), Lateral Occipital Cortex (inferior division), and Superior Parietal Lobule. Coordinates for narcolepsy exhibited stronger connectivity with the Subcallosal Cortex, Frontal Medial Cortex, and Middle Temporal Gyrus (anterior division), while showing weaker connectivity with the Superior Parietal Lobule, Supramarginal Gyrus (anterior division), and Supplementary Motor Cortex. Finally, RBD showed stronger connectivity with the Frontal Medial Cortex, Subcallosal Cortex, and Paracingulate Gyrus, but comparatively weaker involvement with the Inferior Frontal Gyrus (pars triangularis and pars opercularis) and Middle Temporal Gyrus (temporooccipital part) (**Figure 3A**). Network-level analyses further demonstrated that ID-related coordinates were preferentially connected with the somatomotor network (z_mean_ = 1.489, p_spin, FDR_ = 0.005) and the DMN (z_mean_ = 0.308, p_spin, FDR_ = 0.005), but less strongly connected with the frontoparietal network (z_mean_ = -0.171, p_spin, FDR_ = 0.005). OSA-related coordinates showed preferential connectivity with the DMN (z_mean =_ 1.472, p_spin, FDR =_ 0.007) and weaker connectivity with the dorsal attention network (z_mean =_ -0.973, p_spin, FDR =_ 0.007). Narcolepsy-related coordinates showed a similar normative connectivity profile, with weaker connectivity to the salience network (z_mean =_ -1.171, p_spin, FDR =_ 0.009). RBD-related coordinates were preferentially characterized by weaker connectivity with the dorsal attention network (z_mean=_ -1.517, p_spin, FDR =_ 0.014) (**Figure 3B**). Overall, these findings indicate that neuroimaging loci reported across different sleep disorders are embedded within partially distinct large-scale normative functional connectivity architectures. In addition to disorder-specific maps, we performed modality-specific analyses to evaluate connectivity patterns across distinct neuroimaging modalities (see Supplement for details).

**Figure 3.**
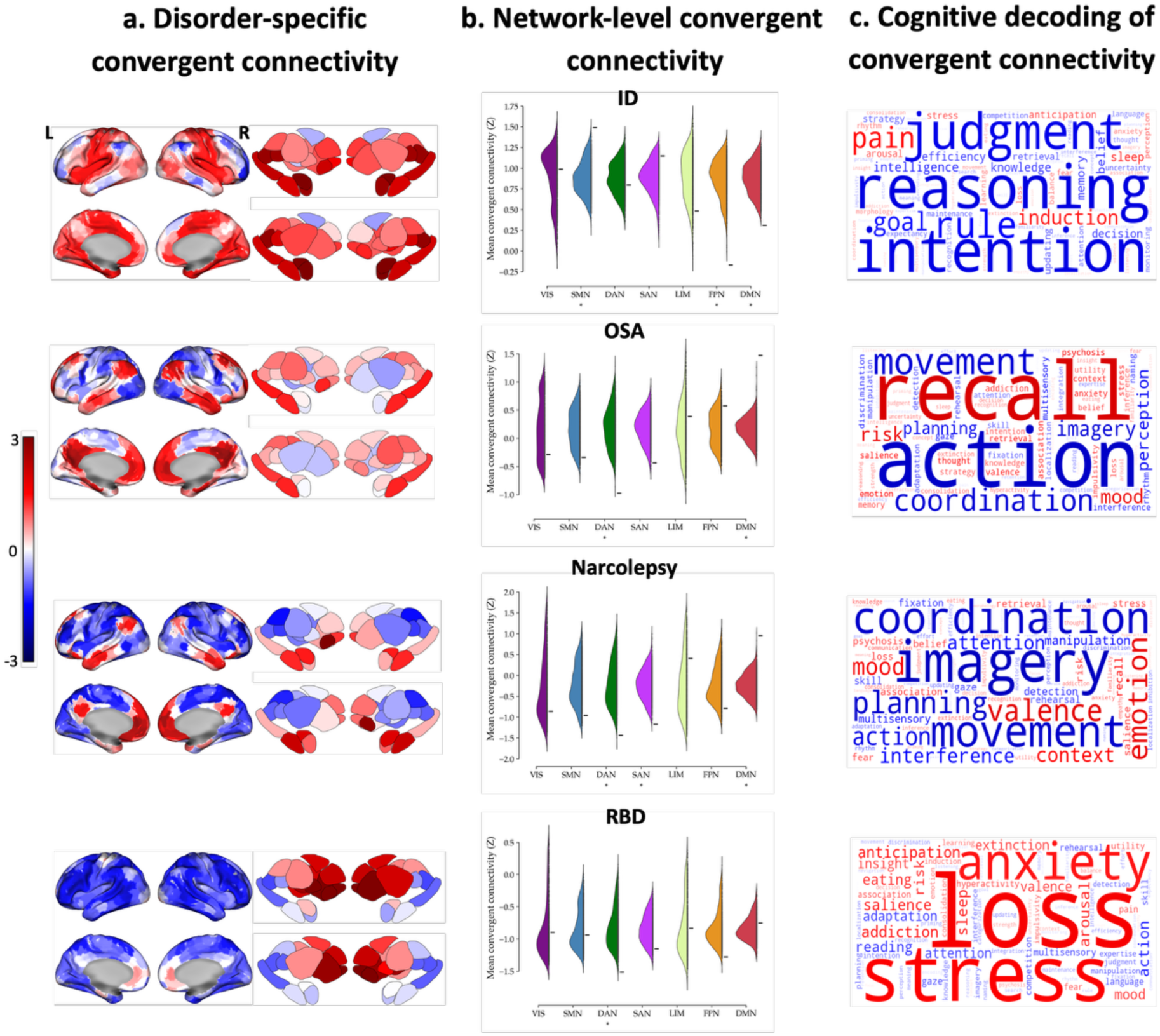
Disorder-specific convergent connectivity maps of sleep disorders. **a)** CCM value distributions across major functional systems show suppression in OSA, narcolepsy, and RBD, contrasting with elevations in insomnia disorder. Within frontoparietal and limbic/midline cortical regions, CCM values were generally increased across sleep disorders, with the notable exception of RBD, which exhibited a divergent downward trend in these areas. **b)** Testing the significance of altered mean spatial association with canonical resting-state networks against a null mean derived from random foci showed that CCM values within the dorsal attention network were significantly lower in obstructive sleep apnea, narcolepsy, RBD. Significance testing revealed an increase in CCM within the default mode network in the sleep disorders except for RBD. CCM values within the somatomotor network were significantly higher, while the frontoparietal network showed a significant decrease in insomnia disorder. Finally, CCM values in narcolepsy within the salience network were significantly lower. Asterisks denote p_spin, FDR_ < 0.05. CCM: convergent connectivity mapping. **c)** To provide a functional interpretation of the convergent connectivity changes observed, the maps were spatially correlated with dynamic consensus term maps from the Neurosynth database. Specifically, a decoding analysis was performed by computing the voxel-wise Spearman correlation between each sleep disorder’s whole-brain convergent maps and Neurosynth’s meta-analytic topic maps. Top terms for each disorder reveal distinct functional themes, with larger text indicating stronger association. ID exhibits cognitive-control related terms; OSA focuses on motor and recall; Narcolepsy emphasizes imagery, planning, and movement; RBD shows high-level cognitive-emotional concepts. *ID: insomnia disorder. OSA: obstructive sleep apnea. RBD: rapid-eye-movement sleep behavioural disorder. VIS: visual network, SMN: somatomotor network, DAN: dorsal attention network, SAN: salience network, LIM: limbic network, FPN: frontoparietal network, DMN: default mode network*.

### Neurotransmitter colocalization

Thus far, we have investigated the brain regions that are connected to the reported abnormalities in sleep disorders using the CCM approach. We next examined the associations between transdiagnostic and disorder-specific CCM maps and PET maps of various neurotransmitter systems. All results were adjusted for multiple comparisons using FDR correction (p_variogram, FDR_ < 0.05). Transdiagnostic CCM values colocalized with serotonin, cholinergic, dopamine, histamine, and glutamate neurotransmitter systems (**Figure 4A**). In disorder-specific analyses, we found an association between the ID’s CCM values and serotonin, norepinephrine, and glutamate systems, whereas the narcolepsy’s CCM values were correlated with the norepinephrine transporter. Additionally, RBD’s CCM values were correlated with serotonin, cholinergic, dopamine, histamine, and GABA systems. In contrast, the OSA’s CCM values showed no significant correlation with any of the measured neurotransmitter systems (**Figure 4B**). Overall, correlations between PET and CCM values indicate that structural and functional brain variations in sleep disorders can potentially be partly explained by variations in neurotransmitter systems.

**Figure 4.**
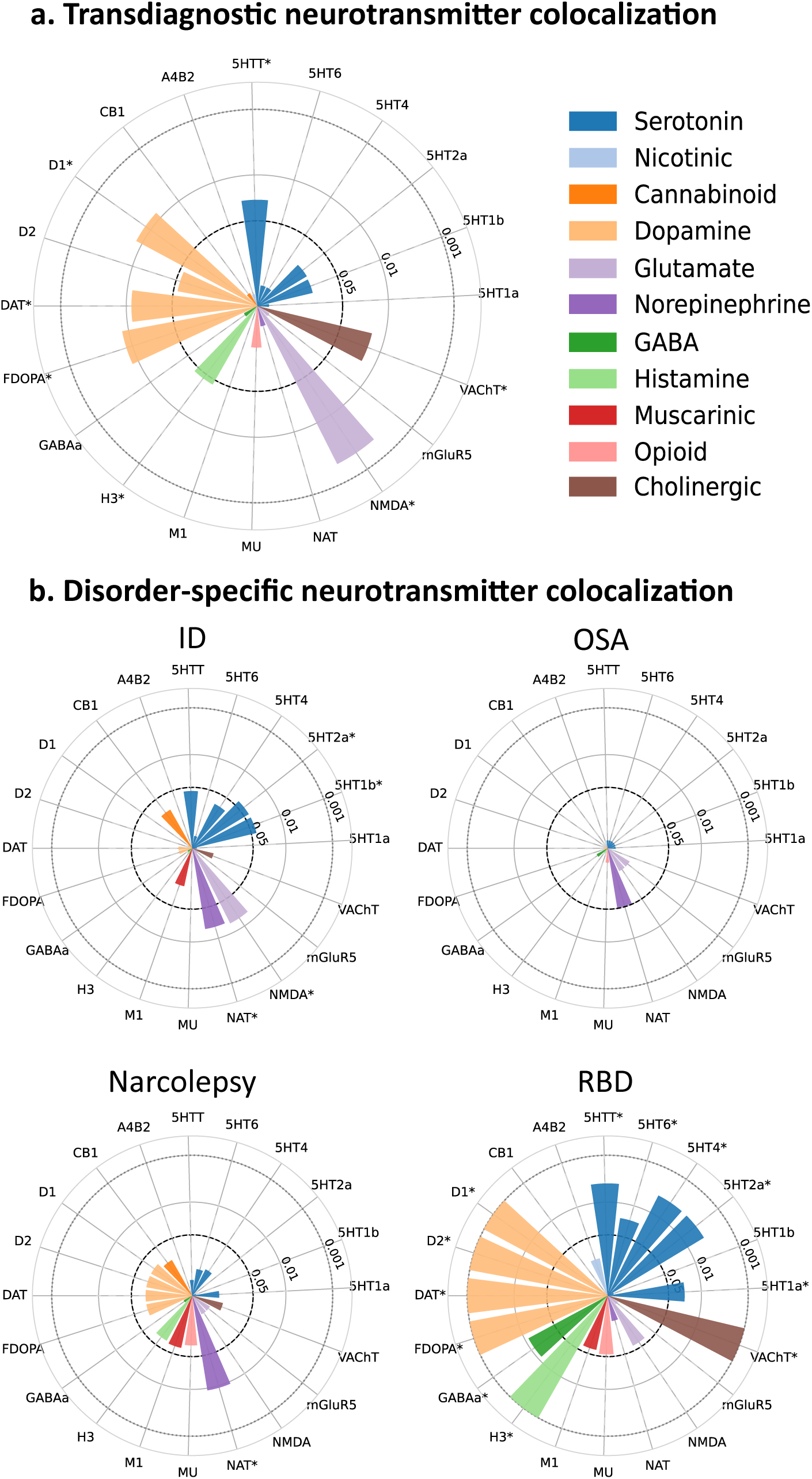
Neurotransmitter co-localization of convergent connectivity maps. (a)The transdiagnostic CCM co-localized significantly with serotonin, glutamate, dopamine, histamine, cholinergic, and histamine systems. (b) The insomnia disorder CCM was related significantly to serotonin, norepinephrine, and glutamate, while the narcolepsy CCM was specifically correlated with the norepinephrine transporter. RBD was significantly co-localized with dopamine, serotonin, cholinergic, histamine, and GABA. Conversely, the obstructive sleep apnea CCM showed no significant correlation with any of the measured neurotransmitter systems. Asterisks denote p_variogram, FDR_ < 0.05. *ID: insomnia disorder. OSA: obstructive sleep apnea. RBD: rapid-eye-movement sleep behavioural disorder*.

### Cellular architectures of the CCM values

We used GSEA to examine the relationship between CCM values and microscale transcriptomic features. The single-cell transcriptomics dataset^42^ used in this study comprises 24 cell types, including layer-specific excitatory neurons, subtype-specific inhibitory neurons, and non-neuronal cell families. After adjusting for gene set size and multiple hypothesis testing (p_FDR_ < 0.05), CHANDELIER and PVALB inhibitory neurons, as well as excitatory neurons within all cortex layers, and non-neuronal cells such as astrocyte and oligodendrocyte precursor cells were enriched for CCM values in the transdiagnostic sleep disorder analysis. Notably, the enriched excitatory and inhibitory neurons were downregulated (NES < 0) and showed an inverse transcriptional association with CCM variations in the transdiagnostic analysis, while non-neuronal cells were upregulated (NES > 0) (**Figure 5A**). Moreover, the cellular enrichment analysis in ID mirrored the transdiagnostic results, whereas no inhibitory neuron was enriched. In contrast, ASTRO, OPC, and VLMC cells, along with VIP, SST-CHODL, and PAX6 inhibitory neurons, were upregulated in OSA, while excitatory neurons (except for L5/6 NP and L6B), CHANDELIER and PVALB inhibitory neurons, and OLIGO cells were downregulated. Additionally, cellular enrichment analysis in narcolepsy reflected the same pattern observed in the transdiagnostic enrichment outcomes, except that SST-CHODL inhibitory neurons were upregulated and OLIGO cells were downregulated. Finally, GSEA results revealed that genes within cell families were enriched for CCM values in RBD, analogous to the transdiagnostic results, except for additional downregulation of LAMP5-LHX6 and SNCG in inhibitory neurons. Taken together, cell systems were enriched for sleep disorders, indicating that cell-related systems underpin sleep disorders (**Figure 5B**).

**Figure 5.**
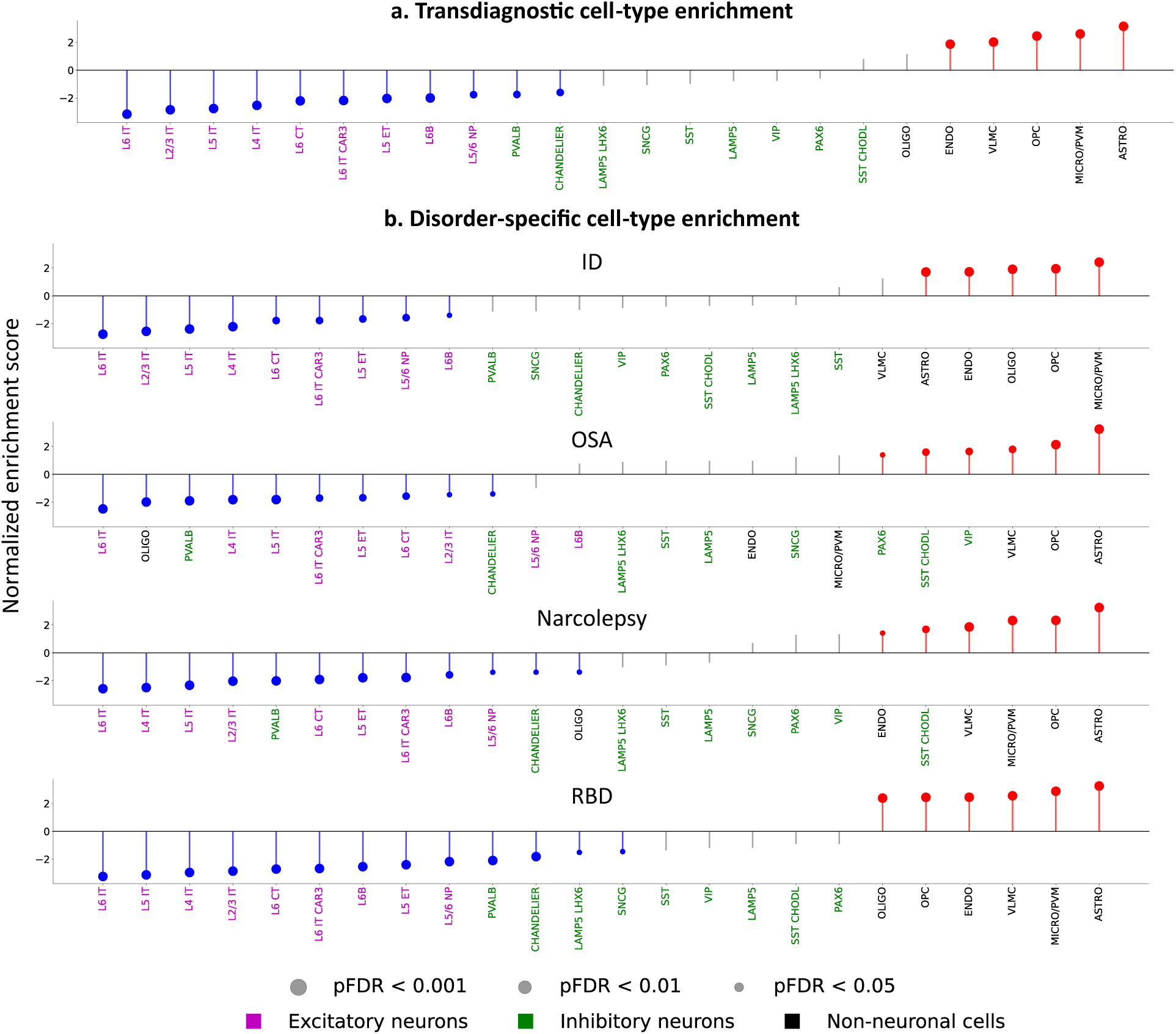
Enriched cell types for convergent connectivity maps. (a) Relationship between transdiagnostic CCM maps and microscale cellular features across 24 distinct classes, including layer-specific excitatory neurons, subtype-specific inhibitory neurons, and non-neuronal families. Significant enrichment (p_FDR_ < 0.05) of CHANDELIER, PVALB, and all cortical excitatory layers in the transdiagnostic analysis, showing inverse transcriptional associations (NES < 0), while astrocytes and OPCs exhibit upregulation (NES > 0). (b) Disorder-specific enrichment patterns: Insomnia mirrors transdiagnostic trends without inhibitory involvement. OSA displays distinct upregulation in ASTRO, OPC, and specific inhibitory subtypes (VIP, SST-CHODL) alongside widespread neuronal downregulation. Narcolepsy and RBD largely align with transdiagnostic outcomes but feature unique deviations in SST-CHODL and LAMP5-LHX6/SNCG populations, respectively. *NES: normalized enrichment score. ID: insomnia disorder. OSA: obstructive sleep apnea. RBD: rapid-eye-movement sleep behavioural disorder*.

## Discussion

The present study provides transdiagnostic and disorder-specific convergent brain connectivity patterns. Similar to *coordinate network mapping*^43^ *and lesion network mapping*^44–46^, CCM assesses whether reported abnormalities converge on specific functional circuits beyond regional overlap, while accounting for connectivity strength and random spatial convergence. Abnormal connectivity in the intrinsic brain network across different sleep disorders, particularly in ID and OSA, has been reported previously.^17,47,48^ Previous studies also highlighted the involvement of the DMN across multiple sleep disorders^15,17,47^, which has been linked to pre-sleep rumination, as well as affective, anxiety, and memory symptoms. Dysfunction of the frontoparietal and dorsal attention networks in acute sleep deprivation and chronic sleep disorders such as ID, OSA, and narcolepsy has been reported^49–53^ and has been associated with impaired cognitive performance, executive control, and attention, common sleep disorder symptoms. However, our CCM findings reflect normative connectivity profiles of reported coordinates derived from healthy participants rather than direct network abnormalities in patients with sleep disorders.

We also examined the microscale colocalization profiles of CCM values and found that receptor and transporter maps of the dopaminergic, noradrenergic, and glutamatergic neurotransmitter systems were associated with CCM values. Indeed, the role of these neurotransmitter systems in sleep is well documented. Elevated catecholamine (i.e., dopaminergic and noradrenergic) activity contributes to hyperarousal, which impairs both sleep initiation and maintenance in patients with ID.^14,30,54^ Norepinephrine oscillation during slow-wave sleep mediates slow vasomotion and, therefore, plays a critical role in glymphatic waste clearance.^55^ In addition to the catecholamine system, glutamatergic overactivity disturbs the balance with the main inhibitory systems (i.e., GABA) and promotes cortical excitation and sleep fragmentation, leading to neurocognitive and affective consequences, as observed in ID and OSA.^14,32^ We also observed an association between CCM values and the dopaminergic system in patients with RBD, which is related to the development of a synucleinopathy.^56–58^ Collectively, long-term imbalances among these neurotransmitter systems disrupt sleep–wake stability and constitute a central underlying pathway in chronic sleep disorders.

Our cellular enrichment analyses further linked CCM values to distinct neuronal and non-neuronal cell populations, suggesting that macroscale network alterations may reflect underlying cellular molecular architecture. Excitatory neurons across cortical layers and PVALB- and CHANDELIER-associated inhibitory neurons showed inverse transcriptional associations with CCM, whereas astrocytes and oligodendrocyte precursor cells showed positive associations. While these findings suggest that transdiagnostic CCM maps are sensitive to cell-type-specific molecular organization, the opposing neuronal associations should not be interpreted as direct evidence of altered excitation-inhibition balance, a conclusion that requires complementary physiological or molecular evidence. The involvement of glial and vascular components further points to their potential roles in sleep homeostasis and in neuroimmune, metabolic, and myelin-related processes relevant to cognitive and affective dysfunction in chronic sleep disorders.^59,60^ Recent evidence suggests that glial cells actively contribute to sleep disruption in Alzheimer disease, including through microglial modulation of arousal circuits independent of amyloid burden^61,62^. Aquaporin-4 water channels of astrocytes’ end feet regulate cerebrospinal– interstitial fluid exchange and waste clearance, particularly during slow-wave sleep.^63,64^ Thus, chronic sleep disorders such as ID and OSA may lead to glial dysfunctions, glymphatic impairment, cognitive decline, and a higher risk of neurodegenerative diseases.^10,28,64–66^ Disorder-specific transcriptomic profiles may further reflect distinct mechanisms, including hyperarousal in ID, hypoxia-related neuroinflammation and vascular remodeling in OSA, and motor disinhibition in RBD.^54,58,67^

## Conclusion

Our findings link macroscale convergent connectivity to microscale neurobiological systems across sleep disorders, suggesting shared neurochemical mechanisms underlying transdiagnostic features. Convergent alterations within the DMN, dorsal attention, and frontoparietal networks were associated with dopaminergic, noradrenergic, and glutamatergic systems, implicating these pathways in sleep disorder pathophysiology. Disorder-specific analyses showed substantial but variable network overlap, while cellular enrichment revealed coordinated changes in excitatory and inhibitory neurons and glial cells, supporting system-level rather than focal abnormalities. Together, these findings highlight the value of multiscale approaches to understanding sleep disorder pathophysiology.

## Data Availability

All data produced in the present study are available upon reasonable request to the authors

## Author Contributions

- Concept and design: Afshani, Saberi, and Tahmasian.
- Mr. Afshani had full access to all study data and is responsible for data integrity and the accuracy of the data analysis.
- Acquisition, analysis, or interpretation of data: Reimann, Afshani, Saberi, and Tahmasian.
- Drafting of the manuscript: Afshani, Saberi, and Tahmasian.
- Critical revision of the manuscript for important intellectual content: Reimann, Chu, Elmenhorst, Müller, Valk, Dukart, Genon, Eickhoff.
- Statistical analysis: Afshani, Saberi.
- Obtained funding: NA.
- Administrative, technical, or material support: Valk, Dukart, Genon, Eickhoff, Tahmasian.
- Supervision: Tahmasian.

## Conflict of Interest Disclosures

Dr. Eickhoff was supported by the Helmholtz Imaging Platform grant (NimRLS, ZT-I-PF-4-010). Dr. Genon was supported by the Deutsche Forschungsgemeinschaft (DFG, GE 2835/2–1, GE 2835/4-1, GE 2835/9-1). Drs. Saberi and Valk were funded by the Max Planck Society. Drs. Saberi and Valk were additionally funded by the Helmholtz Association’s Initiative and Networking Fund under the Helmholtz International Lab grant agreement InterLabs-0015, and the Canada First Research Excellence Fund (CFREF Competition 2, 2015-2016) awarded to the Healthy Brains, Healthy Lives initiative at McGill University, through the Helmholtz International BigBrain Analytics and Learning Laboratory (HIBALL).

## Funding/support

We did not receive any specific funding for this project. The analyses were performed using the Research Centre Jülich infrastructure and the Max Planck Computing and Data Facility.

## Role of the Funder/Sponsor

The funder had no role in the design and conduct of the study; collection, management, analysis, and interpretation of the data; preparation, review, or approval of the manuscript; and decision to submit the manuscript for publication.

## Data Sharing Statement

The peak coordinates of the reported is obtained from a previous publication (doi:10.1001/jamapsychiatry.2025.0488) and is available at https://osf.io/2w5v9/overview. The code for convergent connectivity mapping is available at https://github.com/amnsbr/ccm_tool. The neurotransmitter data are available at https://github.com/amnsbr/antidepressants_meta and PLS analysis we performed using the code available at https://github.com/SarahMorgan/Morphometric_Similarity_SZ.

## Notes

### Competing Interest Statement

The authors have declared no competing interest.

### Author Declarations

Human connectorm project

