## Supplementary information for "Convergent connectivity of sleep disorders and its neurotransmitter and cell enrichment correlates"

### Resting-state fMRI preprocessing and dense functional connectivity computation

For each participant, minimally processed volumetric resting-state fMRI data from the Human Connectome Project were analyzed in MNI152 space. Additional preprocessing included linear detrending, temporal band-pass filtering (0.01-0.08 Hz), spatial smoothing using a 6 mm full-width at half-maximum Gaussian kernel, and voxel-wise standardization of BOLD time series to zero mean and unit variance within each run. Gray matter voxels were defined using the MNI tissue probability map thresholded at >10% gray matter probability, resulting in 211,590 voxels included in subsequent analyses. Preprocessing was conducted separately for each resting-state run prior to concatenation across the four sessions for each participant. For each participant, pairwise Pearson correlation coefficients were computed between the concatenated BOLD time series of all gray matter voxel pairs, yielding a dense resting-state functional connectivity (RSFC) matrix. Correlation coefficients were transformed using Fisher's r-to-z transformation to improve normality prior to averaging across participants. Extreme Fisher-z values were clipped to the range [-4, 4] to reduce the influence of numerical outliers and unstable correlations arising from near-perfect temporal correspondence. Dense RSFC computation was implemented using GPU-accelerated parallel operations with CuPy to enable efficient large-scale matrix computation. Subject-level Fisher-z-transformed RSFC matrices were then averaged elementwise across all 100 participants to generate the final group-averaged dense RSFC matrix used as input for subsequent CCM analyses.

### Meta-Analytic Coordinate Collection and Spatial Distribution

To map the neurostructural and functional landscape of sleep pathology, coordinates reporting significant brain alterations were systematically aggregated from published neuroimaging literature across four major chronic sleep disorders: insomnia disorder (ID), obstructive sleep apnea (OSA), narcolepsy, and rapid eye movement sleep behavior disorder (RBD). The global spatial distribution of all pooled coordinates, as well as those of each individual sleep disorder, were visualized across orthogonal planes (Figure S1).

### Convergent connectivity mapping analysis

CCM analyses were performed to identify convergent functional connectivity patterns associated with reported neuroimaging foci across sleep disorder experiments. For each reported coordinate, the nearest voxel within the gray matter mask was identified in MNI space using Euclidean distance. Coordinates located more than 6 mm from the nearest gray matter voxel were excluded. For each coordinate, a voxelwise resting-state functional connectivity (RSFC) profile was extracted from the group-averaged dense RSFC matrix. Specifically, the RSFC profile corresponded to the connectivity values between the selected voxel and all gray matter voxels included in the dense RSFC matrix. To avoid overrepresentation of experiments reporting larger numbers of foci, RSFC profiles were first averaged within each experiment to generate an experiment-level connectivity map. Experiment-level maps were subsequently combined across studies using sample-size-weighted averaging, such that experiments with larger participant samples contributed proportionally more to the final estimate. To assess statistical significance, the observed CCM map was compared against a voxelwise null distribution generated from 1,000 permutations of randomly sampled foci. For each permutation, the number of sampled foci matched the number reported in each experiment, and random coordinates were constrained to gray matter voxels within the analysis mask. Consequently, statistical inference was based on the empirical distribution of connectivity patterns arising from randomly sampled coordinates under the same sampling framework as the observed data. The same two-level averaging procedure (within-experiment averaging followed by sample-size-weighted averaging across experiments) was applied to the permuted data to preserve the hierarchical structure of the original dataset. CCM values were calculated as z-scores by subtracting the mean of the null distribution from the observed RSFC value and dividing by the standard deviation of the null distribution.

Because the underlying dense RSFC matrix contained exclusively non-negative connectivity values (no anti-correlated connections were observed), statistical maps were thresholded using a one-tailed approach at $z>1.64$ ($p<0.05$). Finally, to evaluate convergence and spatial consistency across different CCM analyses (e.g., across distinct sleep disorder subtypes), spatial correlation analyses were conducted directly between the unthresholded z-score CCM maps. Finally, to evaluate convergence and spatial consistency across different CCM analyses (e.g., across distinct sleep disorder subtypes), spatial correlation analyses were conducted directly between the unthresholded z-score CCM maps.

### Network-Level Analysis and Spatial Permutations

To evaluate the distribution of CCM effects across large-scale functional systems, network-level analyses were performed using the seven canonical resting-state networks (Ref). Spatial spin permutations were used to generate null distributions while preserving the spatial autocorrelation structure of the observed CCM map. Specifically, the CCM map was rotated 1,000 times across the cortical surface, and mean CCM values within each network were recalculated for each permutation. Two-tailed p-values were computed as the proportion of permuted network values exceeding the observed network-average CCM values in absolute magnitude.

### Meta-Analytic Decoding of CCM Maps

To functionally decode the observed CCM maps, we evaluated their spatial similarity against meta-analytic activation probabilistic maps retrieved from Neurosynth (Yarkoni et al., 2011), an automated platform synthesizing functional neuroimaging studies by linking psychological terms to activation coordinates. We downloaded meta-analytic association maps for 123 terms that relate to cognitive processes [Ref]. To establish spatial alignment within a unified anatomical framework, both the empirical z-scored CCM maps and the 123 Neurosynth probabilistic maps were parcellated using a combined scheme comprising the Schaefer 400 cortical atlas (Schaefer et al., 2018) and the Tian subcortical atlas (scale S2; Tian et al., 2020). Spatial correspondence between each CCM map and individual psychological terms was quantified using Spearman's rank correlations across all parcels. The resulting correlation coefficients were rank-ordered to identify the predominant functional domains associated with each CCM pattern. Finally, the top-ranking term-CCM associations were visualized as word clouds, with term font size scaled proportionally to correlation strength to illustrate the cognitive processes most strongly aligned with each convergent connectivity map.

### Spatial Correlation and Inherent Autocorrelation Correction

To quantify the spatial similarity between the transdiagnostic CCM map and each disorder-specific map, we computed pairwise Pearson's correlation coefficients (r) across all CCM maps, generating a comprehensive similarity matrix (Figure S3). Because standard parametric statistical tests artificially inflate significance when applied to spatial neuroimaging data due to inherent spatial autocorrelation (the tendency for nearby voxels to share similar values), a spatially constrained null model was implemented. Specifically, we utilized a variogram-matching approach to generate surrogate brain maps that preserved the exact spatial autocorrelation structure of the empirical data. Voxel-wise statistical significance for the empirical correlation coefficients was determined by benchmarking the true r-values against this permuted null distribution, thresholded at * $p<0.05$ and *** $p<0.001$.

### Leave-One-Disorder-Out Validation Workflow

To evaluate the stability of the transdiagnostic CCM map and ensure that the resulting topography was not driven disproportionately by a single disorder, a leave-one-disorder-out robustness analysis was performed. This involved omitting one specific sleep disorder from the coordinates and recalculating the CCM map for the remaining conditions (Figure S3).

### Neurotransmitter and connectivity colocalization

To investigate the neurochemical relevance of the identified connectivity patterns, we performed a neurotransmitter contextualization analysis by correlating CCM maps with normative PET-derived neurotransmitter receptor and transporter distributions. Neurotransmitter maps representing 11 neurotransmitter systems, including serotonin, dopamine, GABA, glutamate, norepinephrine, histamine, cannabinoid, nicotinic, muscarinic cholinergic, opioid, and related transporter systems, were obtained from previously published PET datasets compiled in the neuromaps framework. These maps reflect regional estimates of receptor density, transporter availability, or binding potential derived from healthy participant cohorts. Both CCM maps and neurotransmitter maps were parcellated using the combined Schaefer et al. cortical atlas and Tian et al.

subcortical atlas to enable region-wise comparison across the whole brain. Following parcellation, parcel values were standardized using Z-score normalization prior to analysis. When multiple PET maps were available for a given neurotransmitter system, maps were combined using sample-size-weighted averaging to reduce tracer-specific variability and emphasize datasets with larger participant cohorts. Associations between neurotransmitter distributions and CCM values were quantified using parcelwise Pearson correlations. Statistical significance was assessed using variogram-based spatial permutations, which generate surrogate brain maps while preserving the spatial autocorrelation structure of the original neurotransmitter distributions. Variogram-preserving null models were selected instead of conventional spin permutations because the analyses included both cortical and subcortical regions, for which spherical surface rotations are less appropriate. Null distributions were generated independently for each neurotransmitter map, and empirical p-values were computed by comparing observed correlations against the corresponding null distributions. Multiple comparisons across neurotransmitter systems were controlled using false discovery rate (FDR) correction, with significance defined as $p_{FDR} < 0.05$.

### Cellular gene set enrichment analysis

Regional microarray gene expression data were obtained from six postmortem adult brains (1 female; age range: 24–57 years, mean ± SD: 42.50 ± 13.38 years) from the Allen Human Brain Atlas (AHBA). Data preprocessing was performed using the abagen toolbox (version 0.1.4). Gene expression data were mapped to a combined atlas comprising the Schaefer et al. cortical atlas and the Tian et al. subcortical atlas, yielding 432 brain regions in MNI space. Microarray probes were first reannotated using updated probe-to-gene mappings, and probes lacking a valid Entrez ID were excluded. Probes were further filtered based on expression intensity relative to background noise, removing probes with expression below background levels in ≥50% of samples across donors. For genes represented by multiple probes, a single representative probe was selected using differential stability, defined as the consistency of regional expression patterns across donors. Differential stability was quantified as the average pairwise Spearman correlation of probe expression profiles across donor brains:

$$\Delta_S(p) = \frac{1}{\binom{N}{2}} \sum_{i=1}^{N-1} \sum_{j=i+1}^{N} \rho\left[B_i(p), B_j(p)\right]$$

where ρ denotes Spearman's rank correlation of probe expression across regions between donor brains $B_i$ and $B_j$, and N represents the total number of donors. The probe with the highest differential stability for each gene was retained for subsequent analyses. Tissue sample coordinates were updated using nonlinear registration implemented with Advanced Normalization Tools (ANTs). To increase spatial coverage, tissue samples were mirrored bilaterally across hemispheres. Samples were assigned to atlas regions if their MNI coordinates fell within 2 mm of a parcel and matched the corresponding hemisphere and gross anatomical division (cortex, subcortex/brainstem, or cerebellum), thereby reducing anatomically implausible assignments. Samples not assigned to any parcel were discarded. For parcels lacking assigned tissue samples, dense interpolated expression maps were generated by assigning each voxel within the parcel to the nearest tissue sample from the same donor. Regional expression estimates were then computed as the distance-weighted average across voxels within the parcel. This interpolation procedure was applied to improve spatial completeness while preserving donor-specific anatomical constraints. To reduce inter-subject variability, expression values were normalized separately for each donor by z-scoring across genes and subsequently across tissue samples. Samples assigned to the same atlas parcel were averaged within each donor and then across donors, yielding regional gene expression matrices containing expression values for 12,506 retained genes. To investigate the association between transcriptomic organization and CCM patterns, Partial Least Squares (PLS) regression was performed using regional CCM values and parcel-wise gene expression profiles. PLS analysis identifies weighted combinations of genes whose spatial expression patterns maximally covary with the observed CCM maps. The stability of gene contributions to the first PLS component was assessed using bootstrap resampling with 10,000 iterations, and genes were ranked according to their bootstrap-estimated weights. Stable positively and negatively weighted gene sets were subsequently submitted to Gene Set Enrichment Analysis (GSEA) to identify enriched cellular systems using a recently published single-cell RNA sequencing reference dataset. Statistical significance for enrichment analyses was determined using false discovery rate (FDR) correction, with significance defined as $p_{FDR} < 0.05$.

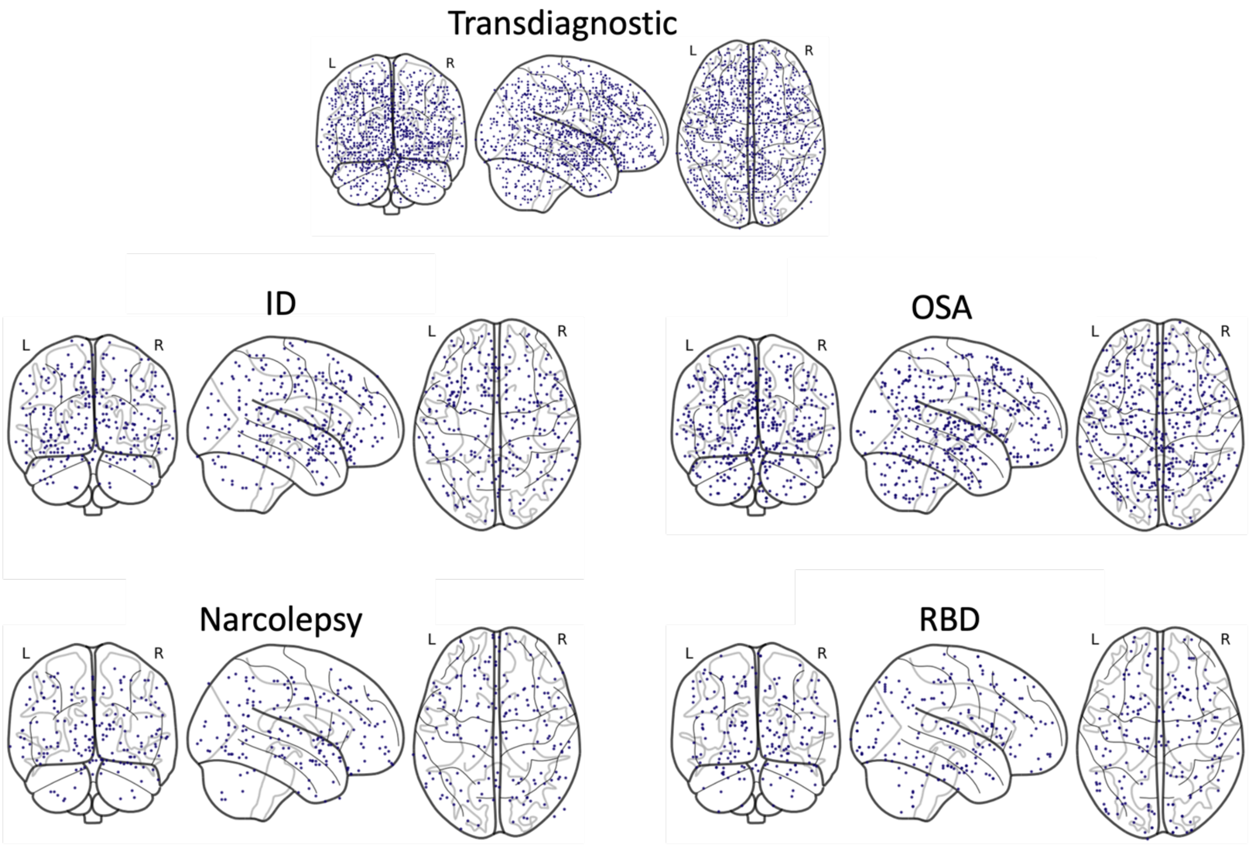


**Figure S1. Spatial distribution of neuroimaging meta-analysis coordinates across sleep disorders.** The top panel displays the global distribution of all aggregated coordinates across the entire study. The middle and bottom panels illustrate the specific spatial distribution of stereotaxic coordinates (blue dots) derived from published neuroimaging literature for individual sleep disorders. Each condition is displayed across three anatomical perspectives: coronal/posterior (left), sagittal/lateral (middle), and axial/superior (right). L, left hemisphere; R, right hemisphere. ID: insomnia disorder. OSA: obstructive sleep apnea. RBD: rapid-eye-movement sleep behavioural disorder.

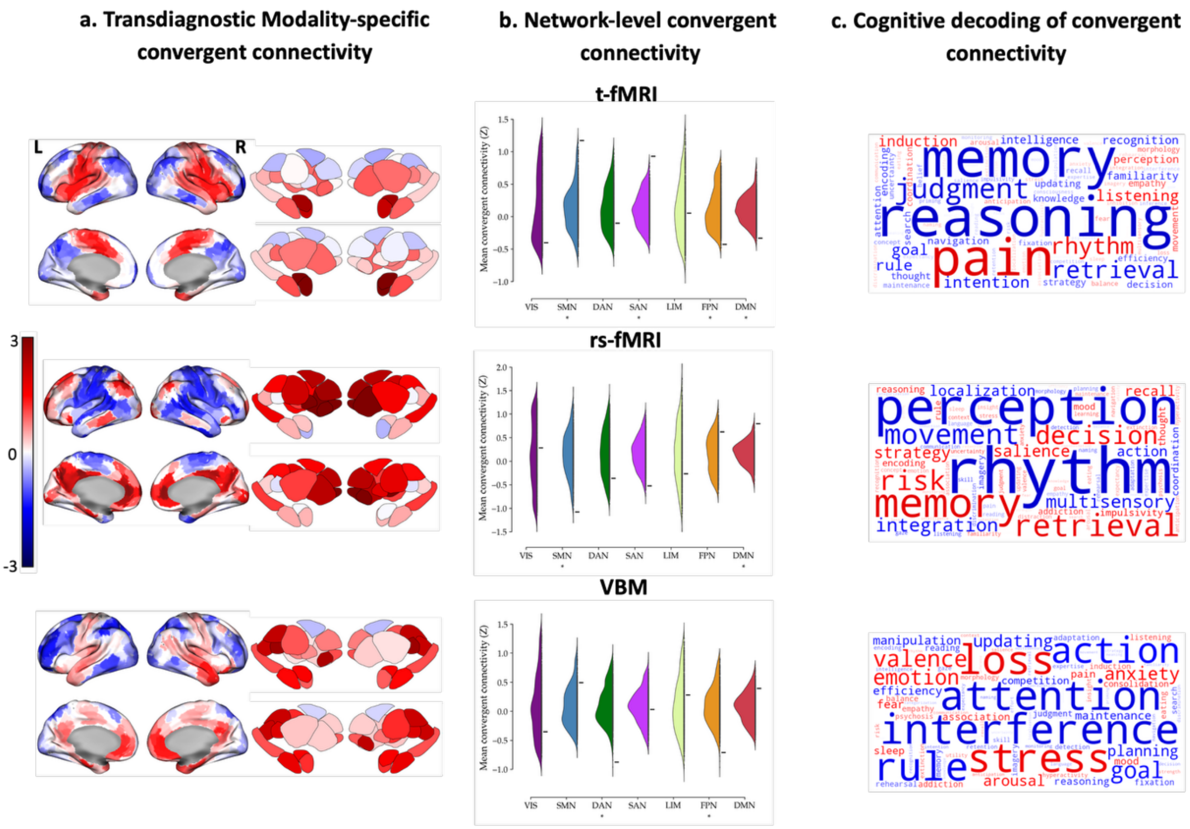


**Figure S2. Modality-specific convergent connectivity mapping. a)** Surface projections illustrate cortical and subcortical CCM value distributions across major functional systems for each neuroimaging modality. **b)** Spatial association testing against permutation-derived null models reveals network-level shifts, with asterisks denoting significant alterations across canonical resting-state networks (p spin ,FDR<0.05). **c)** Cognitive term decoding via spatial correlation with meta-analytic functional maps from the Neurosynth database highlights the predominant functional themes associated with each modality's topography, where word size reflects association strength. Abbreviations: CCM, convergent connectivity mapping; VIS, visual network; SMN, somatomotor network; DAN, dorsal attention network; SAN, salience network; LIM, limbic network; FPN, frontoparietal network; DMN, default mode network.

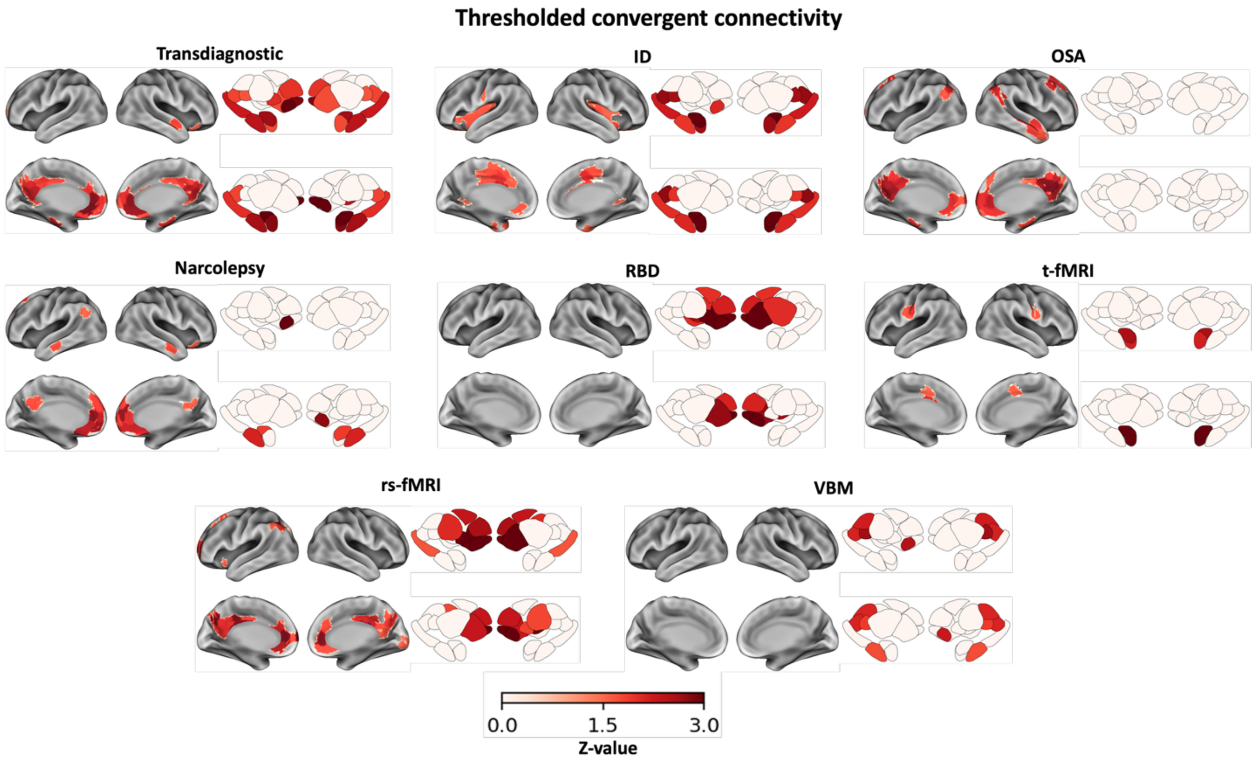


**Figure S3.** Thresholded convergent connectivity mapping (CCM) patterns across shared, disorder-specific, and modality-specific analyses. Cortical and subcortical surface renderings display spatial distributions of CCM values thresholded at Z>1.64 (corresponding to a one-tailed p<0.05). The panel illustrates shared connectivity effects (top left), along with distinct disorder-specific topographies (e.g., insomnia disorder, obstructive sleep apnea, narcolepsy, and REM sleep behavior disorder) and modality-specific patterns across neuroimaging modalities (structural, functional, and diffusion-weighted imaging). The color bar indicates standardized Z-scores, with darker red regions representing stronger convergence exceeding the significance threshold. Abbreviation: CCM, convergent connectivity mapping.

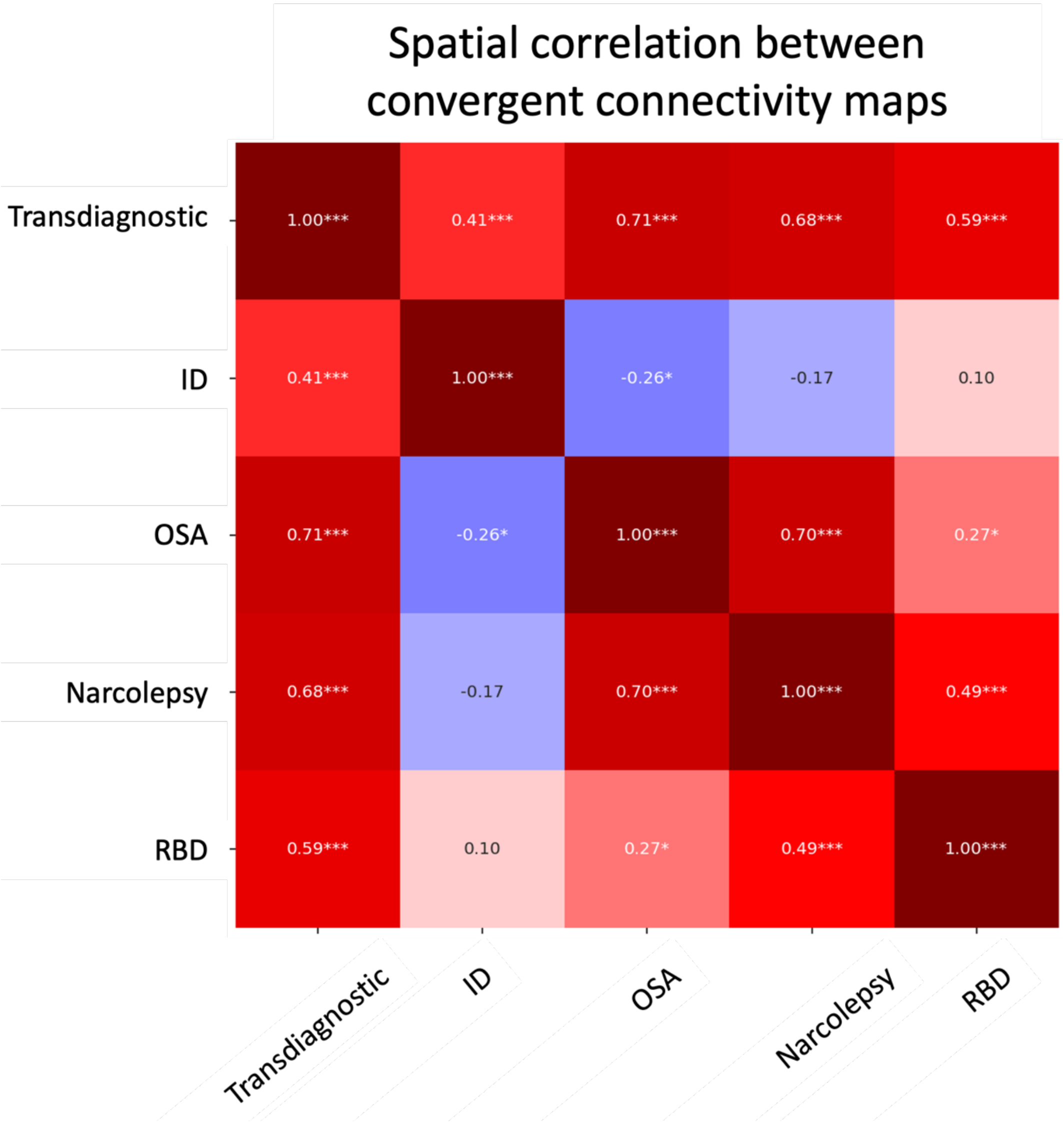


**Figure S4. Spatial correlation matrix of transdiagnostic and individual sleep disorder CCM maps.** The heatmap displays the pairwise spatial similarity (Pearson's correlation coefficients) between the transdiagnostic convergent connectivity mapping map and individual maps. The color scale denotes the strength and direction of the correlation, ranging from blue (negative correlation) to dark red (positive correlation). Statistical significance was determined using a spatially constrained null model derived from the variogram matching test to account for spatial autocorrelation (* $p<0.05$, *** $p<0.001$). The color bar shows Pearson correlation coefficients. CCM: convergent connectivity mapping; ID: insomnia disorder; Narco: narcolepsy; OSA: obstructive sleep apnea; RBD: rapid eye movement sleep behavior disorder.

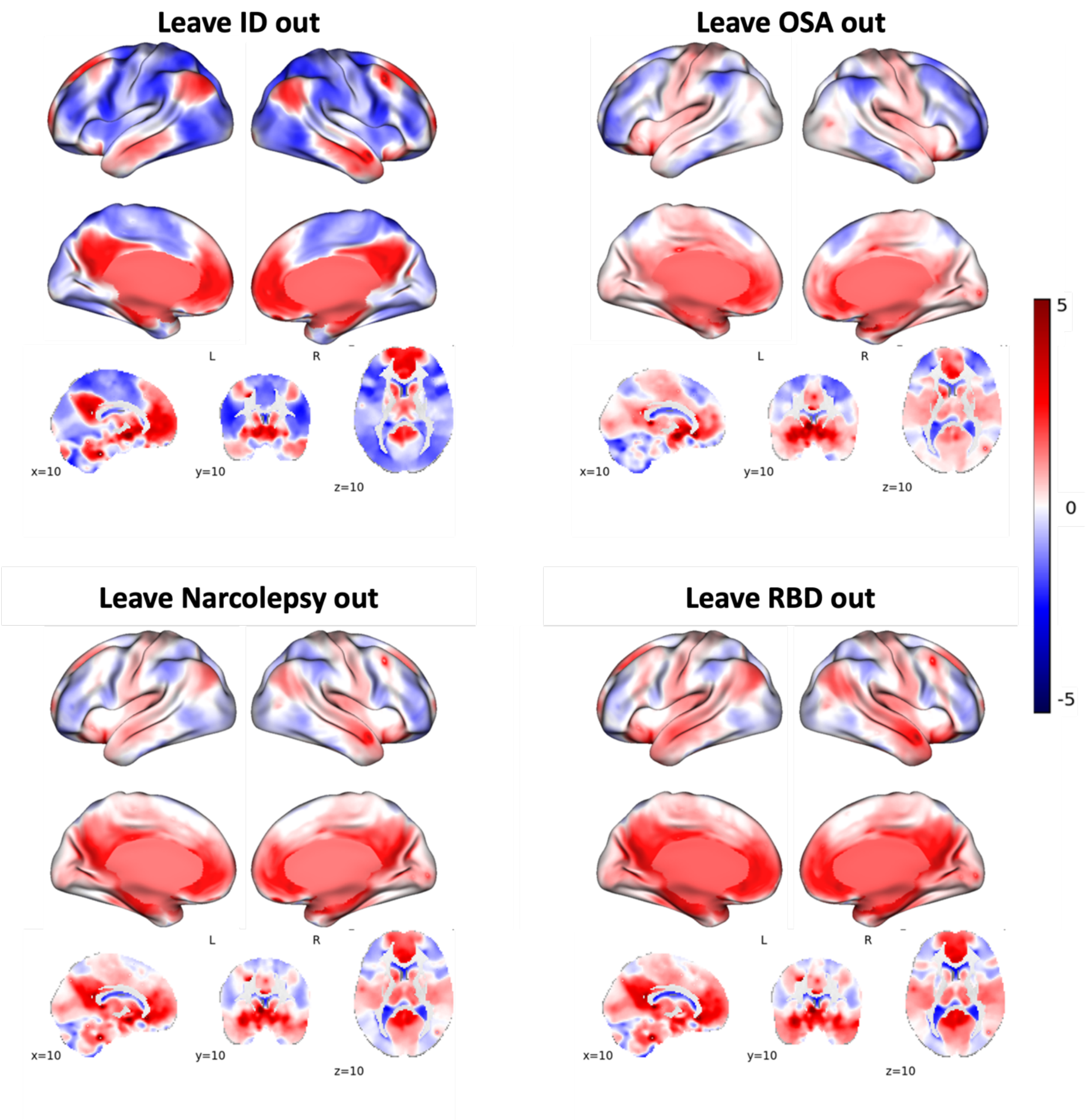


**Figure S5. Structural distribution of transdiagnostic CCM maps under individual sleep disorder exclusion.** The panels display the remaining cortical and subcortical co-activation topographies when systematically omitting one specific sleep disorder from the Convergent Connectivity Mapping analysis. The color bar shows z-values. L, left hemisphere; R, right hemisphere. ID: insomnia disorder. OSA: obstructive sleep apnea. RBD: rapid-eye-movement sleep behavioural disorder.
